# Development of a Metastatic Uveal Melanoma Prognostic risk Score (MUMPS) for use in patients receiving immune checkpoint inhibitors

**DOI:** 10.1101/2021.02.28.21252611

**Authors:** Deirdre Kelly, April A. N. Rose, Thiago Pimentel Muniz, David Hogg, Marcus O. Butler, Samuel D. Saibil, Ian King, Zaid Saeed Kamil, Danny Ghazarian, Kendra Ross, Marco Iafolla, Daniel V. Araujo, John Waldron, Normand Laperriere, Hatem Krema, Anna Spreafico

## Abstract

**Background:** Metastatic uveal melanoma (mUM) is a rare disease for which no systemic therapy has demonstrated overall survival (OS) benefit. There are no robust data on prognostic factors for OS in patients with mUM treated with immune checkpoint inhibitors (ICI).

Retrospective and non-randomized prospective studies have reported response rates of 0-37% for anti-PD1/L1 +/-anti-CTLA4 ICI in mUM, indicating a potential benefit only in a subset of patients. This study evaluates the characteristics associated with ICI benefit in patients with mUM.

**Methods:** We performed a single-center retrospective cohort study of patients with mUM who received anti-PD1/L1 +/-anti-CTLA4 ICI between 2014–2019. Clinical and genomic characteristics were collected from chart review. Treatment response and clinical progression were determined by physician assessment. Multivariable Cox regression models and Kaplan-Meier log-rank tests were used to assess differences in clinical progression-free survival (cPFS) and OS between groups and to identify clinical variables associated with ICI outcomes.

**Results:** We identified 71 mUM patients who received 75 lines of ICI therapy. Of these, 54 received anti-PD1/L1 alone, and 21 received anti-PD1/L1 + anti-CTLA4. Patient characteristics were: 53% female, 48% were 65 or older, 72% received one or fewer lines of prior therapy. Within our cohort, 53% of patients had developed stage IV disease < 2 years after their initial diagnosis. Bone metastases were present in 12% of patients. For the entire cohort, the median cPFS was 2.7 months and median OS was 10.0 months. In multivariable analyses for both cPFS and OS, the following variables were associated with good prognosis: ≥ 2yrs from initial diagnosis to stage IV (n=25), LDH <1.5xULN (n=45), and absence of bone metastases (n=66). We developed a **M**etastatic **U**veal **M**elanoma **P**rognostic risk **S**core (**MUMPS**). Patients were divided into 3 MUMPS risk groups based on the number of the above-mentioned prognostic variables: Poor risk (0-1), Intermediate risk (2) and Good risk (3). Good risk patients experienced longer cPFS (6.0 months) and OS (34.5 months) than patients with intermediate (2.3 months cPFS, 9.4 months OS) and poor risk disease (1.8 months cPFS, 3.9 months OS); P<0.0001.

**Conclusion:** We developed a MUMPS risk score, based on retrospective data, that is comprised of 3 readily available clinical variables (time to stage IV diagnosis, presence of bone metastases, and LDH). This MUMPS risk score has potential prognostic value. Further validation in independent datasets is warranted to determine the role of this MUMPS risk score in selecting ICI treatment management for mUM.

## Background

Uveal melanoma is the most common primary intraocular malignant tumor in adults. Up to 50% of patients diagnosed with uveal melanoma will develop metastatic disease[1]. Historical one-year survival rates of metastatic uveal melanoma (mUM) range from 13% to 15%[2]. Uveal melanoma disseminates hematogenously, and 80–90% of patients with mUM will present with the liver as the first site of metastasis [3, 4].

No systemic therapy has thus far been reported to demonstrate survival benefit for patients with mUM, and the National Comprehensive Cancer Network (NCCN) guidelines prioritize clinical trial enrollment [5]. Standard practice for managing mUM, outside of clinical trial enrollment, involves systemic therapy or liver-directed therapies, including surgery when the disease is mainly limited to the liver [6-8]. Systemic therapy approaches include the same treatments that have demonstrated efficacy in cutaneous melanoma, including anti-programmed cell death protein 1 (anti-PD1) and anti-cytotoxic T-lymphocyte antigen-4 (anti-CTLA4) immune checkpoint inhibitors (ICI) [9]. However, patients with uveal melanoma have been excluded from many of the randomized phase 3 clinical trials that have demonstrated the efficacy of ICI in cutaneous melanoma. Retrospective studies with anti-PD1/L1 monotherapy in mUM have reported objective response rates (ORR) of less than 5%, median PFS of 3 months (0.75–6.75) with a median OS of 5 months (1–16) [10, 11]. In retrospective and prospective studies of patients receiving anti-PD1/L1 + anti-CTLA4, ORR ranges from 11 to 18% with median PFS of 3-5.5 months and OS of 15-20 months [12-15].Recent work published on UM patients that received expanded access to anti-PD1/L1 + anti-CTLA4 found that, at a median follow up of 17.8 months, median OS was nor reached[16].

The combination of anti-PD1/L1 + anti-CTLA4 is often considered the preferred regimen for metastatic uveal melanoma due to the higher ORR [14, 17, 18]. However, anti-PD1/L1 and anti-CTLA4 ICI remains a highly toxic regimen, with severe toxicities reported in 40-64% of patients [13, 17, 19].Grade 3-4 toxicities frequently lead to early treatment discontinuation and/or long-lasting toxicities that can significantly impact patients’ quality of life.

In the absence of randomized phase III evidence for OS benefit from anti-PD1/L1 ICI regimens in patients with mUM, there is a clinical need to define predictive biomarkers of ICI response that could assist in the optimal treatment selection for patients and to identify high-risk prognostic features in this patient population to guide clinical management. This retrospective analysis aims to evaluate the clinical, biochemical and molecular characteristics with prognostic and predictive value in a cohort of mUM patients treated with anti-PD1/L1 ICI either as monotherapy or in combination with anti-CTLA4 ICI.

## Materials and Methods

### Study Design

This is a retrospective single-center cohort study. This study was approved by the Research Ethics Board at the University Health Network - Princess Margaret Cancer Centre (REB# 19-5186) and was conducted in accordance with the principles of Good Clinical Practice, the provisions of the Declaration of Helsinki, and other applicable local regulations.

### Study Population

The Princess Margaret University Health Network (PM-UHN) Tumor Immunotherapy Program (TIP) and melanoma referral databases were used to identify patients with uveal melanoma who received anti-PD1/L1-based ICI (either alone or in combination with anti-CTLA4) as palliative treatment for advanced disease. Eligible patients were defined as those with a diagnosis of stage IV uveal melanoma receiving therapy with anti-PD1/L1 ICI with or without an anti-CTLA4 ICI. Patients could have received prior therapies in any treatment line. Cases were collected from January 2014 and December 2019 with baseline imaging and follow-up data. Demographics, treatment parameters, clinical genomic data and clinical outcomes of interest were extracted from the original patient records and merged into a central database before analysis.

### Data Collection and Treatment Outcomes

Clinical data retrieved from electronic medical records included: age, sex, sites of metastatic disease at treatment initiation, size of metastases, baseline Eastern Cooperative Oncology Group (ECOG) performance status, anti-PD1/L1-based regimen used, toxicity, and previous systemic therapies. Serum lactate dehydrogenase (LDH) and neutrophil to lymphocyte ratio (NLR) at the time of treatment initiation were collected and analyzed for their prognostic value.

Genomic data were available for a subset of patients and were collected retrospectively. This included information from multiplex ligation-dependent probe amplification (MLPA) testing, and Next Generation Sequencing (NGS). Impact Genetics MLPA testing was done at the time of primary tumor treatment and used to estimate the probability of metastatic death after treatment of uveal melanoma[20]. MLPA comprises a set of probes, each hybridizing to a specific genomic sequence where chromosome 3 loss and chromosome 8q gain predict poor prognosis, and chromosome 6p gain is associated with improved outcomes and decreased risk of metastasizing [20-22]. Within our cohort, NGS was done on recurrent, metastatic UM using either the UHN Melanoma NGS Panel Version 1.1 or the Oncomine Comprehensive Assay v3 (ThermoFisher). The UHN Melanoma NGS Panel examined exonic coding regions of *BAP1, BRAF, CDK4, CDK6, CDKN2A, GNA11, GNAQ, KIT, NRAS, EIF1AX*, and *SF3B1*. This methodology employs SureSelect Target Enrichment hybrid capture followed by paired-end sequencing on the Illumina sequencing platform. Variant calls were generated using the UHN Clinical Laboratory Genetics custom bioinformatics pipeline with alignment to genome build GRCh37/hg19, and assessed using Cartagenia Bench Lab NGS v5. The Oncomine Assay is a targeted, next-generation sequencing (NGS) assay that enables the detection of relevant SNVs, indels, CNVs and gene fusions in 161 cancer-related genes. All of the above genes on the UHN Melanoma NGS panel, with the exception of EIF1AX are included in this assay. Sequencing was performed on the Ion S5 XL System, with data analysis using Ion Reporter 5.12 (ThermoFisher). Variant annotations were obtained from OncoKB when variants were present in the OncoKB database[23].

The best radiologic response to treatment was assessed independently by study investigators (DK and AANR). Patients were typically restaged with CT scans or MRI every 12 weeks as part of routine clinical care. Best overall response was calculated using baseline and follow-up imaging assessments made by the study investigators (DK, AANR). Tumor response (TR) was defined as radiological evidence of tumor shrinkage of any lesion in the absence of any new growth, as reported by the radiologist. Progressive disease (PD) was defined as significant tumor growth, according to the treating physician assessment of radiologic imaging. Stable disease (SD) was defined as any response that did not meet the criteria for either TR or PD. RECIST criteria were not employed. Clinical progression was based on physician documentation of disease progression or death in the electronic medical record, according to either clinical or radiologic parameters of progression. Clinical progression-free survival (cPFS) was calculated from the date of initiation of ICI treatment to documented clinical progression, death or last follow-up, defined as date of death or last clinic visit. Study data-lock occurred on September 8, 2020. Patients who were alive without clinical progression at last follow-up were censored. Overall survival (OS) was calculated from the date of initiation of anti-PD1/L1 ICI to death or last follow-up. Reason for treatment discontinuation was also collected. Data were extracted by a first investigator (DK) and reviewed by a second one (AANR). Discordances in treatment response evaluation were resolved by consensus.

### Data Analysis and Statistics

Differences in patient characteristics between treatment groups were examined using Fisher’s exact test. Student’s T-test was used to assess statistically significant differences in the number of cycles of therapy received between treatment groups. Univariable and multivariable Cox models were used to evaluate differences in OS and cPFS. Cox regression models were used to calculate Hazard ratios (HR) with 95% confidence intervals (CI) and P-values for survival analyses. We included all clinical variables in the initial multivariable model. Non-significant variables were removed by backward step-wise selection. The final multivariable models for cPFS or OS included only those variables that were associated with P<0.10 in multivariable analysis. Kaplan-Meier Survival curves were compared with the log-rank test. All statistical analyses were performed using Stata v12.

## Results

### Patient characteristics

A total of 71 patients with mUM who received 75 lines of anti-PD1/L1 +/-anti-CTLA4 ICI for metastatic disease at our institution were identified (Supplementary Figure 1). In total, 54 patients received anti-PD1/L1 monotherapy, and 21 received anti-PD1/L1 + anti-CTLA4 (**Table 1**). Thirty-eight patients (51%) were naïve to systemic treatment and received ICI as first-line systemic therapy. The median age at the time of ICI was 64 (range 34-89) years, and 48% of patients were 65 or older at the time of treatment initiation. Within the study population, 40 (53%) patients were female, and the majority (69%), were ECOG PS 1 at the time of ICI (**Table 1**). Body mass index (BMI) was 25 or higher in 68% of patients. Serum LDH was elevated over 1.5 times the upper limit of normal in 40% of patients at baseline. Bone metastases were present in 12%, whereas 96% had liver metastases. Other baseline characteristics are listed in **Table 1**. Patients who received anti-PD1/L1 + anti-CTLA4 therapy were more likely to have a BMI of 25 or higher than patients who received anti-PD1/L1 monotherapy (P=0.054). There was a trend towards younger age in the group that received anti-PD1/L1 + anti-CTLA4, but this was not statistically significant (P=0.130). The remaining variables were not significantly different between those who received anti-PD1/L1 monotherapy and anti-PD1/L1 +/-anti-CTLA4.

**Table 1:**
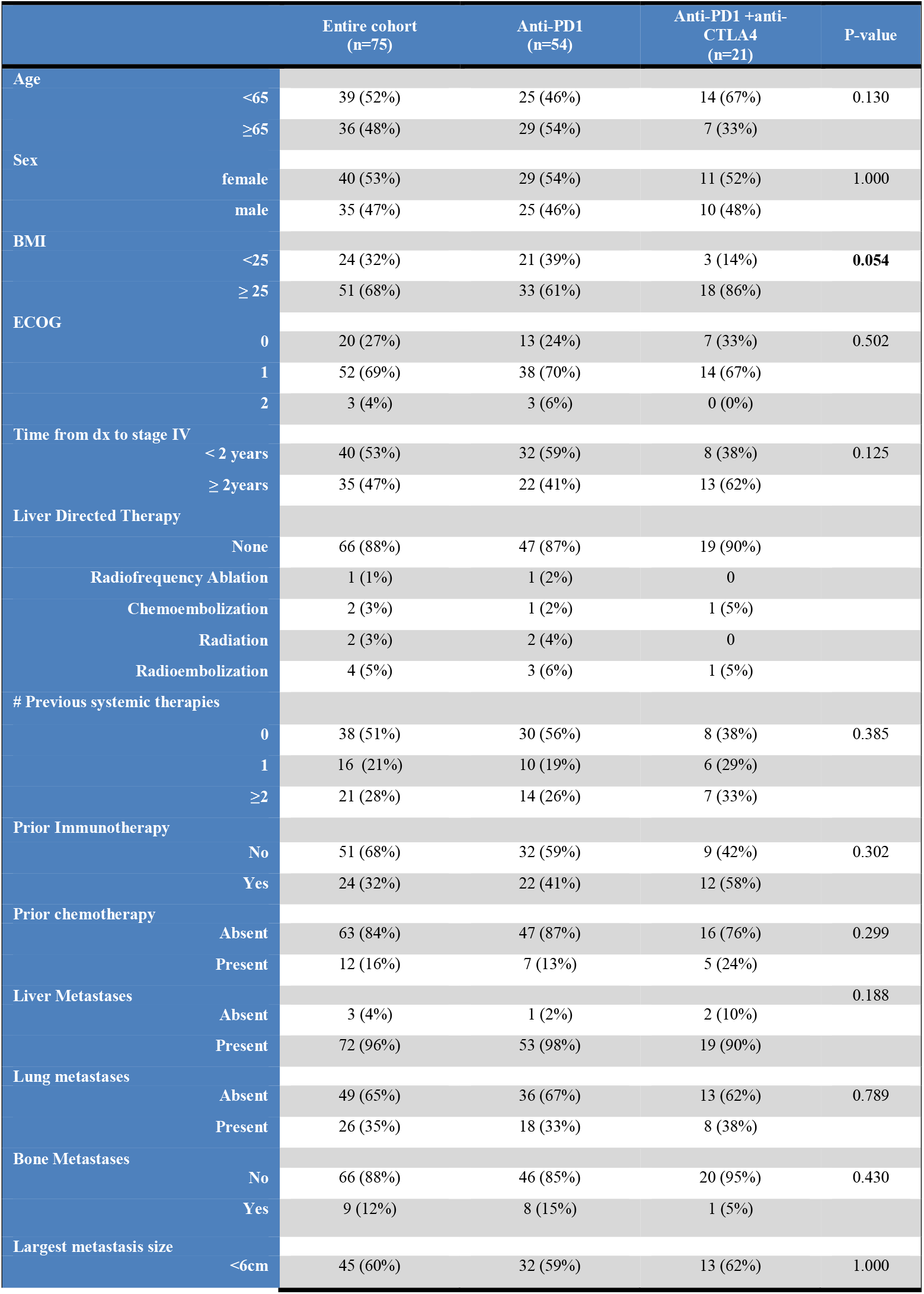
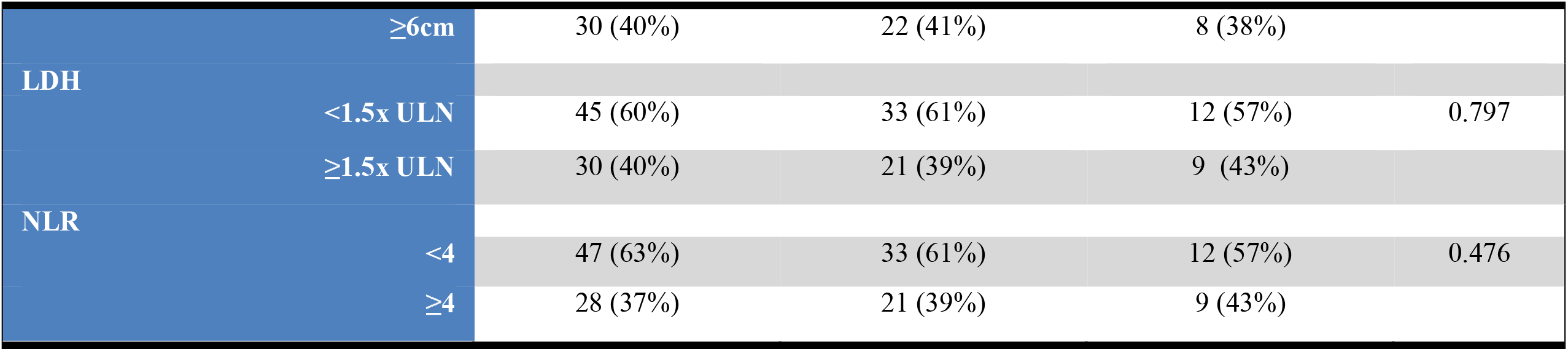
Clinical characteristics of 71mUM patients who received 75 lines of anti-PD1/L1 +/- anti-CTLA4 ICI.

**Figure 1:**
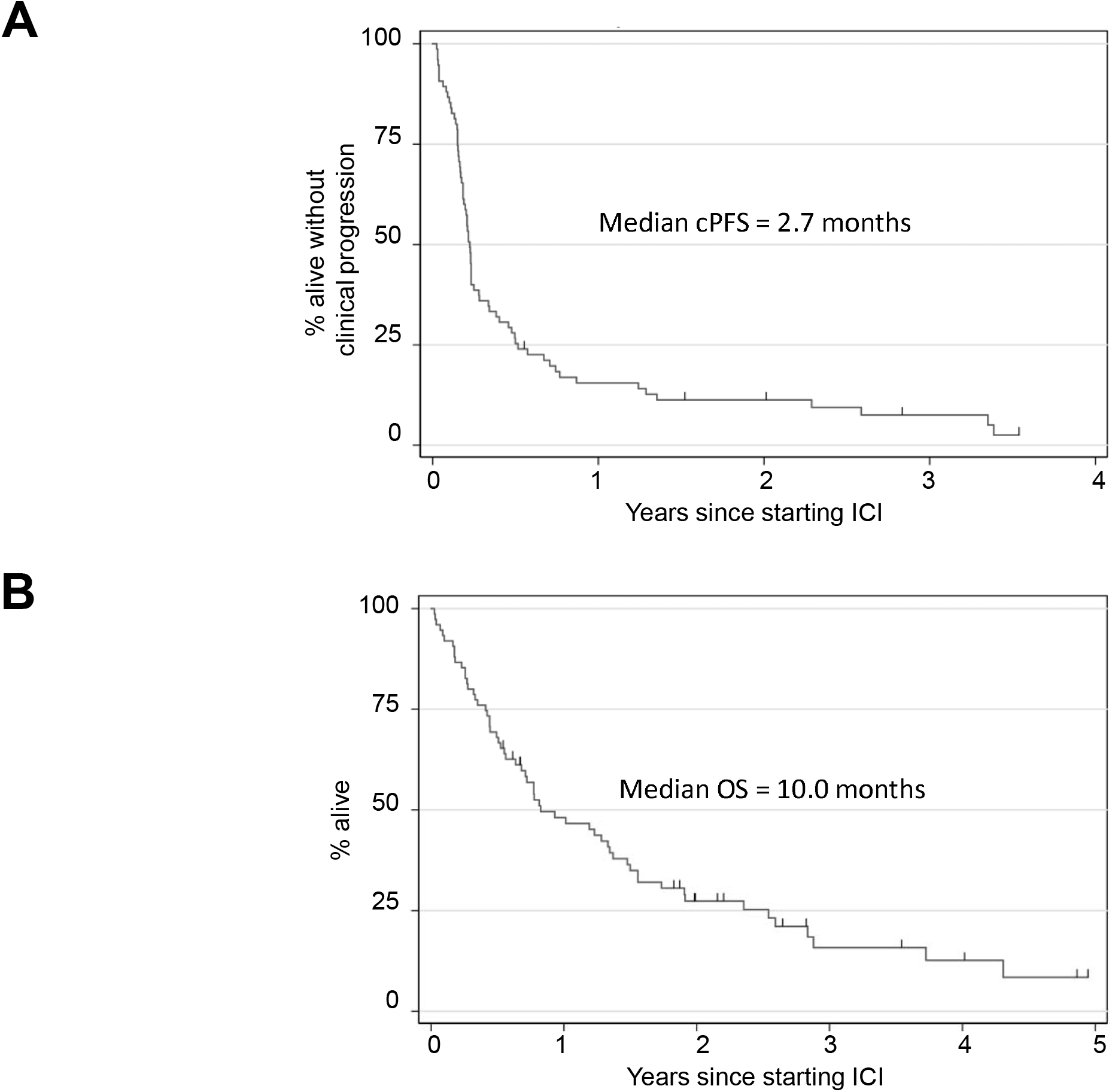
Survival of metastatic uveal melanoma (mUM) patients who received anti-PD1 +/-anti-CTLA4 ICI. **(A)** Clinical progression-free survival and **(B)** overall survival of the entire cohort of 71 mUM patients who received 75 lines of therapy. Tick marks indicate censored patients.

### ICI treatment duration and toxicity

In our entire cohort, 72 (96%) treatments were discontinued at the time of last follow-up. There were no treatment-related deaths that occurred during the observation period. The majority, 61 (81%), discontinued treatment due to disease progression, and 9 (12%) discontinued therapy due to toxicity. Two patients, both of whom received anti-PD1/L1 monotherapy, had their care transferred to an outside hospital and were lost to follow-up and 3 patients were still receiving ICI treatment (**Table S1**). Of the 21 patients who received anti-PD1/L1 + anti-CTLA4, 17 (81%) received high dose anti-CTLA4 (3mg/kg) with low dose (1mg/kg) anti-PD1/L1 Q3W followed by anti-PD1/L1 monotherapy at standard doses, and 4 (19%) received low dose anti-CTLA4 (1mg/kg) combined with standard dose anti-PD1/L1 (2-3mg/kg) Q3W followed by anti-PD1/L1 monotherapy at standard doses (**Table S2**). Only 7 (33%) patients who started anti-PD1/L1 + anti-CTLA4 received all 4 pre-planned doses (**Table S1**). The majority of patients (57%) who started anti-PD1/L1 + anti-CTLA4 treatment did not go on to receive any anti-PD1/L1 monotherapy. The median number of treatment cycles that included anti-PD1 monotherapy given at standard doses was higher (4 cycles, range 1-38) than it was for patients who received anti-PD1/L1 + anti-CTLA4(2 cycles, range 0-29, P=0.0079). Patients who received anti-PD1/L1 + anti-CTLA4 were more likely to have discontinued therapy due to toxicity (4/21; 19%) than patients who received anti-PD1/L1 monotherapy (5/54, 9%), but this difference was not statistically significant (**Table S1**).

### ICI treatment outcomes

For the entire cohort of patients, the median cPFS was 2.7 months (**Figure 1a**), and the median OS was 10.0 months (**Figure 1b**). Of these patients, 10 (13%) had a treatment response (TR), 15 (20%) had SD and 49 (65%) had PD as best response. One patient was lost in follow-up before they could be evaluated for treatment response. Treatment responses were observed in 11% of patients who received anti-PD1/L1 monotherapy and 19% of patients who received anti-PD1/L1 + anti-CTLA4. The differences in the rate of treatment response were not statistically significant (P = 0.589) (**Figure 2a**). ICI regimen type was also not associated with significant differences in cPFS (**Figure 2b**) or OS (**Figure 2c**).

**Figure 2:**
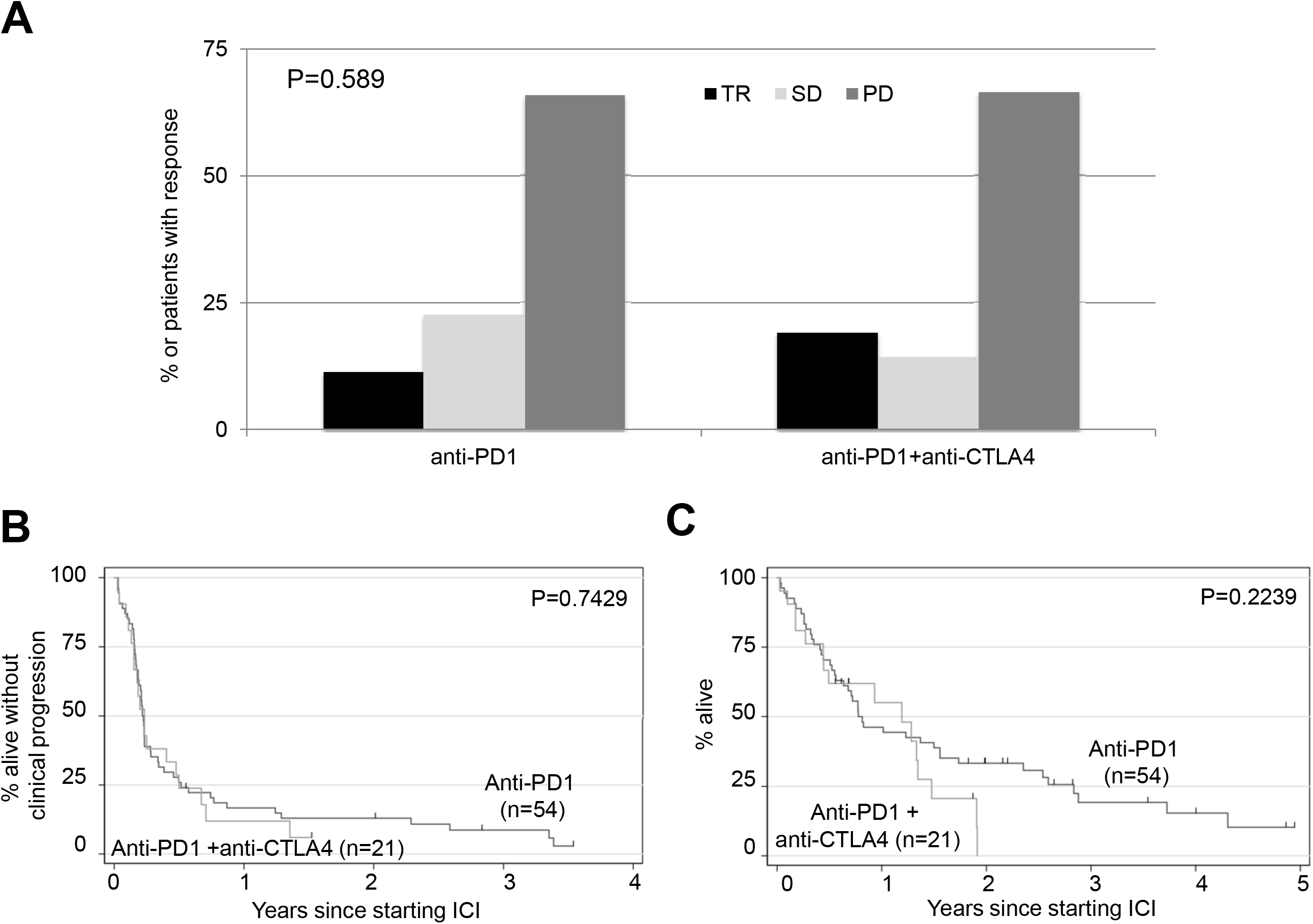
Treatment response and survival outcomes according to anti-PD1 regimen. **(A)** Treatment response according to whether patients received anti-PD1 monotherapy (n=54) or anti-PD1 and anti-CTLA4 (n=21). TR = tumor response, SD = stable disease, PD = progressive disease. The difference in distribution of treatment responses according to ICI regimen type was assessed with a fisher exact test (P=0.589). **(B)** Clinical progression-free survival and **(C)** overall survival according to anti-PD1 ICI regimen. Log-rank test P= 0.7429 (cPFS); P=0.2239 (OS). Tick marks indicate censored patients.

### Genomic analyses of study population

In this study, 16 patients (21%) had Impact Genetics chromosomal testing on primary tumor samples, and 50 (67%) received NGS testing on metastatic specimens. Molecular characteristics were heterogeneous across this study and differed between the group of patients who received anti-PD1/L1 alone and those who received anti-PD1/L1 + anti-CTLA4. Impact testing was more likely to have been performed among patients who received anti-PD1/L1 + anti-CTLA (33%) vs. anti-PD/L1 monotherapy (17%), but this difference was not statistically significant. Patients who received anti-PD1/L1 + anti-CTLA4 were more likely to have chromosome 6p disomy (33%) compared to those who received monotherapy (9%, P=0.049), although only a minority of tumors was tested in both groups. As expected, all tested mUM tumors were *BRAF* and *NRAS* wild-type. Amongst patients who received anti-PD1/L1 monotherapy, 32 (59%) had tumors tested for NGS; *GNA11* and *GNAQ* mutations were present in 24% and 21% of patients, respectively. Patients who were treated with anti-PD1/L1 + anti-CTLA4 were more likely to have had NGS (86%, P=0.032). In this group, *GNA11* and *GNAQ* mutations were present in 19% and 52% of patients, respectively. As such, *GNAQ* mutations were more common in this group (P=0.024). Similarly, *BAP1* mutations were more commonly identified amongst patients who received anti-PD1/L1 + anti-CTLA4 (54%) compared to those who received anti-PD1/L1 monotherapy (15%, P=0.003). *SF3B1* mutations were observed with similar frequencies in both treatment groups (10% vs. 7%, respectively) (**Table S3**). *GNA11* and *GNAQ* mutations were mutually exclusive in our population and each individual gene was examined in relation to clinical outcomes with no single gene significantly associated with cPFS and OS (data not shown).

### Identification of clinical variables associated with survival

We employed univariate and multivariate cox-regression analyses to assess whether any clinical variables were associated with cPFS or OS in our cohort of mUM patients treated with anti-PD1/L1 +/-anti-CTLA4 ICI. Clinical variables that were associated with shorter cPFS in multivariate analyses were: time from the initial diagnosis to metastatic disease ≤2 years (P = 0.001), presence of bone metastasis (P □ = □ 0.086), LDH≥ 1.5 times the upper limit of normal (xULN) (P = 0.002) and neutrophil-lymphocyte ratio (NLR) ≥ 4 (P = 0.005) (**Table 2**). Clinical variables associated with shorter OS in multivariate analysis were: time from initial diagnosis to metastatic disease of less than 2 years (P < 0.001), presence of bone metastasis (P = 0.016), metastasis with a diameter ≥ 6cm (P = 0.006), LDH≥ 1.5x ULN (P < 0.001) (**Table 3**).

### Prognostic value of MUMPS risk score

We sought to develop a prognostic risk score that would have clinical utility for prospectively identifying a cohort of mUM patients that were most likely to have good survival outcomes. Our **M**etastatic **U**veal **M**elanoma **P**rognostic risk **S**core (MUMPS) included only those “good risk” clinical variables that were associated (P<0.10) with both improved cPFS (**Table 2**) and improved OS (**Table 3**) in multivariable analyses. These variables were: i) time from the initial diagnosis to metastatic disease ≥ 2 years, ii) absence of bone metastases, iii) LDH ≤ 1.5 x ULN. Patients were divided into risk groups according to the number of MUMPS risk factors that they possessed: Good risk (3); Intermediate risk (2) and poor risk (0-1). We found that 29%, 51% and 20% of patients belonged to the good, intermediate, and poor risk groups, respectively (**Figure 3a, Table S4**). Patients classified as having good risk also had significantly longer median cPFS (6.0 months) compared to those with intermediate (2.3 months) or poor risk (1.8 months) (P=0.0001, **Figure 3c**). Good risk patients experienced significantly longer median OS (34.5 months) compared to patients with intermediate (9.4 months) or poor risk scores (3.9 months) (P<0.0001, **Figure 3d**).

**Figure 3:**
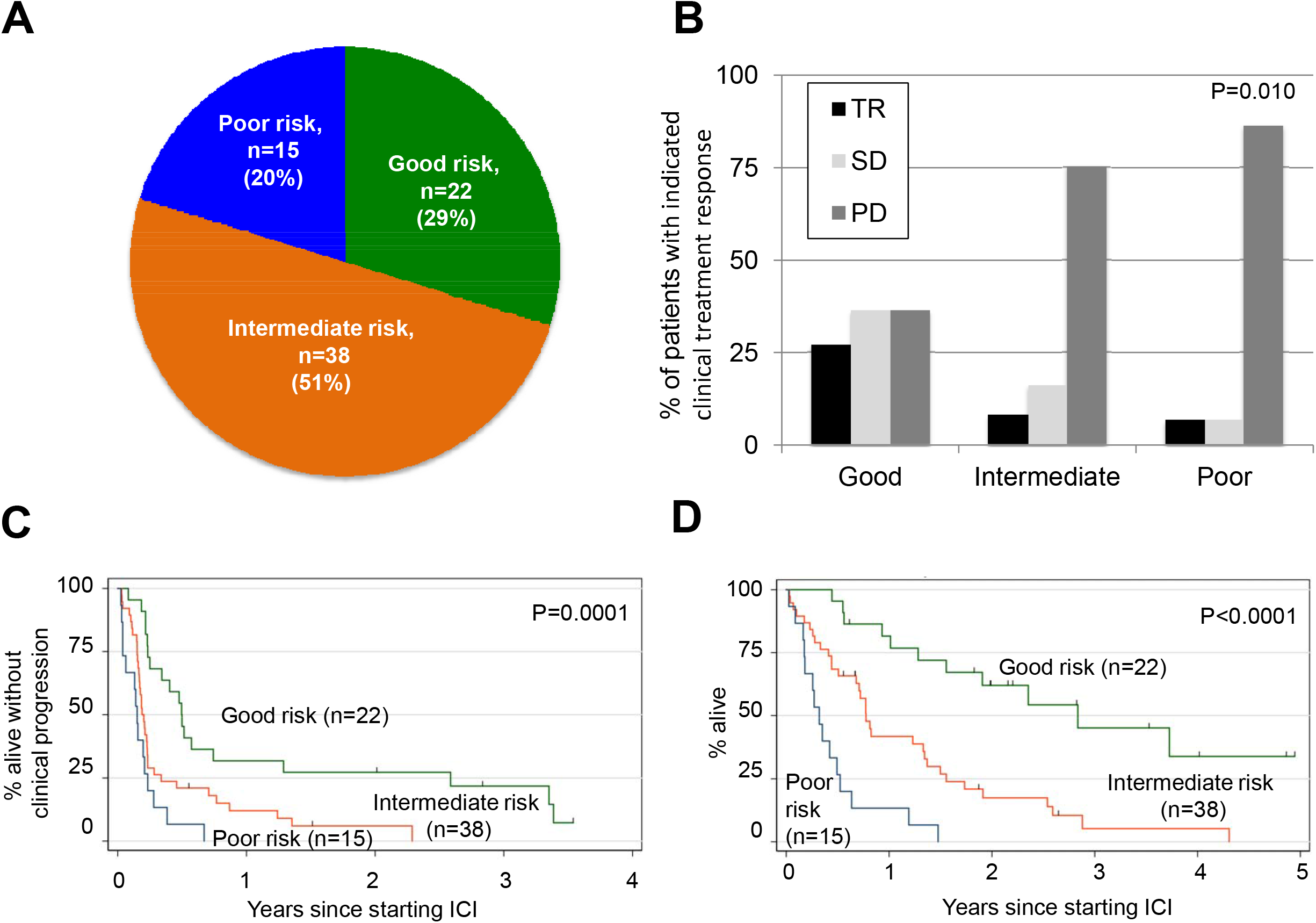
Association between Metastatic Uveal Melanoma Prognostic risk Score (MUMPS) groups and clinical outcomes. Patients were categorized based on the number of MUMPS good risk factors they possessed. Good risk factors were: absence of bone metastases, ≥ 2 years from initial diagnosis to stage IV disease, LDH <1.5x ULN. A) The percentage of patients classified as having good risk (MUMPS = 3) intermediate risk (MUMPS = 2) and poor risk (MUMPS = 0-1) are indicated. B) Percent incidence of clinical treatment responses to ICI, according to MUMPS risk group, are indicated; Fisher’s exact test, P=0.010 C) cPFS according to MUMPS risk group. D) OS according to MUMPS risk group. Log-rank test P=0.0001 (cPFS); P<0.0001 (OS). Tick marks indicate censored patients. Good risk (n=22) = green line, Intermediate risk (n=38) = orange line, Poor risk (n=15) = blue line. TR = tumor response, SD = stable disease, PD = progressive disease.

### Predictive value of MUMPS risk score

Next, we assessed whether the MUMPS risk groups could predict benefit from either anti-PD1/L1 monotherapy or anti-PD1/L1 + anti-CTLA4. Patients classified as having a good risk based on their MUMPS risk category were more likely to have achieved TR or SD as best clinical treatment response and least likely to have achieved PD as best clinical treatment response compared to patients with intermediate or poor-risk prognosis, P=0.010 (**Figure 3b**). For patients with a MUMPS good risk score, there was a non-significant trend (P=0.0750) toward longer cPFS among those who received anti-PD1/L1 monotherapy compared to anti-PD1/L1 + anti-CTLA4(**Figure 4a**). MUMPS good-risk patients treated with anti-PD1/L1 monotherapy experienced significantly longer OS (45.3 months) compared to patients with a good prognosis who received anti-PD1/L1 + anti-CTLA4 (15.6 months) P=0.0042 (**Figure 4b**). There was no significant difference in cPFS (p=0.456) (**Figure 4c**) or OS (p=0.756) (**Figure 4d**) according to anti-PD1/L1 treatment regimen, among patients with intermediate or poor-risk prognosis.

**Figure 4:**
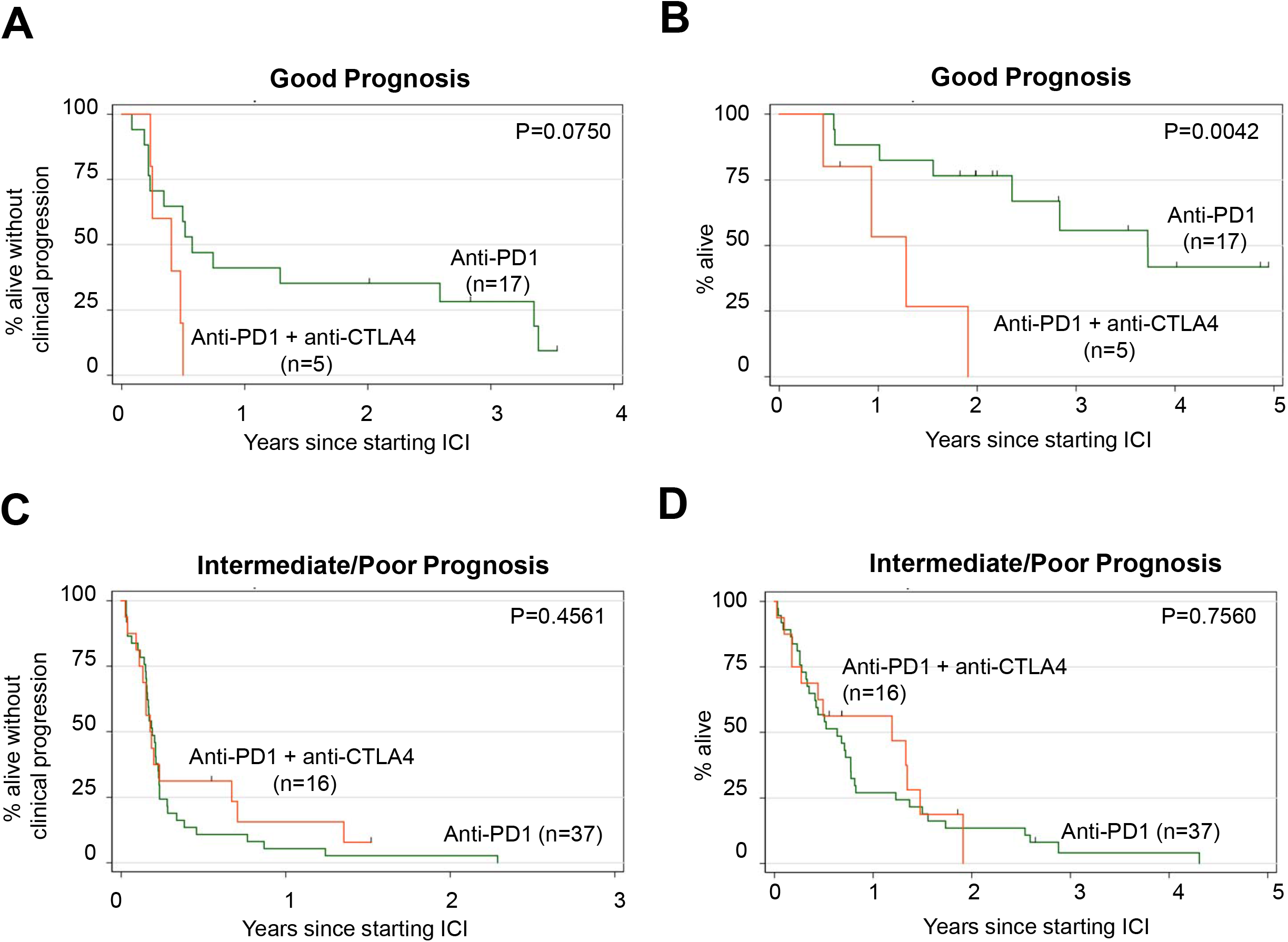
Survival according to MUMPS risk group and ICI regimen. Survival outcomes of patients with good prognosis (MUMPS = 3) were stratified according to the anti-PD1 regimen they received: anti-PD1 monotherapy (n=17), anti-PD1 and anti-CTLA4 (n=5). cPFS is shown in A) and OS is shown in B. Log-rank test P=0.0750 (cPFS); P=0.0042 (OS). Survival outcomes of patients with intermediate (MUMPS = 2) or poor prognosis (MUMPS = 0-1) were stratified according to the anti-PD1 regimen they received: anti-PD1 monotherapy (n=37), anti-PD1 and anti-CTLA4 (n=16). cPFS is shown in A) and OS is shown in B. Log-rank test P=0.4561 (cPFS); P=0.7560 (OS).

## Discussion

In contrast to cutaneous melanoma, ICIs have limited activity in mUM, and the standard of care ICI regimen in this setting has not formally been established. Indeed, we observed no significant differences in treatment outcomes when comparing anti-PD1/L1 monotherapy to anti-PD1/L1 + anti-CTLA4 in our entire cohort. Enrollment in clinical trials is an NCCN guideline recommendation for patients with mUM, and prognostic models are needed to better understand the natural history of this rare disease, to stratify patients enrolling in clinical trials, and to facilitate treatment discussions and decisions for clinicians and patients.

In this study, we describe a retrospective cohort of 71 mUM patients who received 75 lines of anti-PD1/L1 +/-anti-CTLA4 ICI, and we undertake a comprehensive analysis of clinical variables associated with outcome. The median cPFS (2.7 months) and OS (10.0 months) that we observed in this study were very similar to established benchmarks for mUM patients receiving systemic therapy (3.3 and 10.2 months, respectively)[24]. This suggests that our cohort is reasonably representative of the greater mUM patient population. Based on multivariable analyses, we developed a prognostic risk score (MUMPS) that is associated with clinically meaningful differences in survival outcomes among a cohort of mUM patients.

The **M**etastatic **U**veal **M**elanoma **P**rognostic risk **S**core (**MUMPS**) developed in this study is composed of one laboratory value and two clinical variables that are associated with good outcomes. These variables are readily accessible for assessment by treating physicians and include: ≥ 2 years from initial diagnosis to stage IV disease, absence of bone metastases and LDH < 1.5x ULN. Each of these clinical variables has previously been identified in multiple independent studies of survival outcomes in metastatic uveal melanoma patients, and each have demonstrated prognostic value, irrespective of treatment type[12, 24, 25]. The clinical prognostic factors in our MUMPS risk model may be reflective of patients with less aggressive tumor biology. Indeed, short time to relapse and elevated LDH are factors that have been previously reported in other disease sites and may be due to increased tumor burden, aggressive tumor biology, and paraneoplastic processes [26-29]. Bone metastases are also known to be associated with poor prognosis in many other metastatic cancers, including non-small cell lung cancer, urothelial cancer, head and neck cancer, and renal cell carcinoma (RCC) [30-33]. The reasons for this association are not fully understood, but may reflect an increase in paraneoplastic processes such as hypercalcemia, or poor quality of life and/or functional status among patients with painful bone metastases that limit further systemic therapy interventions or clinical trial enrolment.

Risk stratification and employment of prognostic criteria have proven beneficial for treatment selection in other disease sites. For example, the International Metastatic Renal Cell Cancer Database Consortium (IMDC) criteria [28] is a clinically useful prognostic risk score that is comprised of clinical variables associated with OS in metastatic RCC. Interestingly, although this IMDC prognostic score was derived from a group of patients who all received anti-angiogenic targeted therapies, it also has clinical utility in predicting the therapeutic benefit of immune checkpoint inhibitors [34]. Similarly, we found that our MUMPS prognostic score may also have predictive value. We showed that, in our cohort, patients with MUMPS good risk derived more benefit from anti-PD1/PDL1 alone vs. anti-PD1/L1 + anti-CTLA4. However, amongst patients with intermediate and poor risk MUMPS scores, there was no clear benefit associated with either anti-PD1/L1 regimen. These observations require independent prospective validation prior to clinical implementation.

Based on predominantly retrospective data and single-arm prospective studies, anti-PD1/L1 + anti-CTLA4 is generally the preferred regimen due to relatively higher rates of reported objective responses compared to anti-PD1/L1 monotherapy in cross-study comparisons <u>[14, 17]</u>. Our data challenges the belief that anti-PD1/L1 + anti-CTLA4 should be the preferred systemic ICI for all mUM patients. Indeed, we observed no significant differences in the clinical response rate, cPFS or OS amongst patients who received anti-PD1/L1 + anti-CTLA4 versus anti-PD1/L1 monotherapy. In our cohort, patients with good prognostic MUMPS score survived longer with anti-PD1 monotherapy than with anti-PD1/L1 + anti-CTLA4. This result was surprising, but there could be a number of biologic rationale that would explain this observation. Patients who received anti-PD1/L1 + anti-CTLA4 were more likely to discontinue treatment due to toxicity and received less cycles of treatment that contained standard dose anti-PD1/L1 ICIs. It is also possible that increased receipt of subsequent immunosuppressive therapy due to immune related adverse effects in the anti-PD1/L1 + anti-CTLA4 group may mitigate the anti-tumor effects of ICI. In our study, the majority of patients who received anti-PD1/L1 + anti-CTLA4 either progressed early or developed treatment related AEs and did not ever go on to receive to anti-PD1/L1 monotherapy maintenance. Responses to anti-PD1/L1 are not as long lasting in mUM compared to cutaneous melanoma[11, 35], and in mUM even complete responses are not necessarily durable for the long term [19]. We speculate that increased duration of exposure to full dose anti-PD1 may be particularly beneficial in mUM, however this would require further study in independent datasets. Despite this observation, the reported response rates of 12-18% among patients who received anti-PD1 + anti-CTLA4 in prospective trials makes anti-PD1 + anti-CTLA4 a compelling therapeutic option[17, 36]. It would be of interest to assess whether anti-PD1 + anti-CTLA4 with lower dose anti-CTLA4 might represent an alternative option to maximize the therapeutic benefit with less dose-limiting toxicity than standard anti-PD1 + anti-CTLA4 dosing regimens. Indeed, low dose anti-CTLA4 (1mg/kg) with full dose anti-PD1 -either 3 mg/kg or 2mg/kg has been investigated for the treatment of cutaneous melanoma in Checkmate-511 [37] or KEYNOTE-029 [38]. These combination regimens with low dose anti-CTLA4 appear to have similar efficacy to combination ICI with high dose anti-CTLA4 and have been reported to have less serious adverse events than high dose ICI anti-PD1 + anti-CTLA4, and may result in less treatment discontinuation due to toxicity and less requirement for immunosuppressive therapy.

In cutaneous melanoma, it is well established that responses can be maintained for a long period of time after ICI treatment discontinuation[39]. However, the fundamental underlying biology of uveal melanoma is different from cutaneous melanoma[40], and we do not yet know if ICI responses are maintained after treatment discontinuation in this population. Indeed, continuous treatment with anti-PD1 ICI beyond a year is associated with longer OS compared to 1-year fixed duration of treatment in non-small cell lung cancer[41]. In a separate, prospective study of 35 mUM patients who received anti-PD1 + anti-CTLA4, discontinuing treatment due to toxicity was not associated with longer PFS [17]. More research is needed to better understand the consequences of holding or discontinuing ICI treatments due to serious immune-mediated adverse events in uveal melanoma, as this may influence further anti-PD1 + anti-CTLA4 drug development, as well as clinicians’ readiness to re-institute ICI after an adverse event. It could be speculated that for some patients with good prognostic features (i.e., those with MUMPS good risk disease) the risk of discontinuing or holding treatment early, and increased use of immunosuppressive therapy due to increased toxicity with anti-PD1 + anti-CTLA4 vs. anti-PD1 monotherapy may out-weigh the benefits of potentially being more likely to obtain an objective response. Additional studies are required to validate this hypothesis.

Current experimental strategies focus on various single-agent and combination strategies targeting: Melanocyte protein PMEL**/**GP100, Procaspase-Activating Compound 1, NTRK Inhibitors, LAG-3 Antibodies, Focal Adhesion Kinase (FAK) Inhibitors, Protein kinase C Inhibitors and MEK Inhibitors (**Table S5**)[42]. Radioembolization and immunoembolization continue to be studied in combination with anti-PD1 +/-anti-CTLA4. Alternate immune-modulating approaches using autologous tumor-infiltrating lymphocytes (TILS) and T cell receptor (TCR) activation are being explored. Autologous TILS have shown evidence of tumor regression in mUM as well as responses in patients refractory to ICI and IMCgp100, a bispecific molecule that targets the T-cell receptor has shown antitumor activity and remains in development [43, 44].

This study is subject to a number of limitations. This was a retrospective study that analyzed outcomes for patients treated in a single academic center in Canada; therefore, the results may not be generalizable to the broader community. Although this is one of the largest cohort studies to be published for ICI treated mUM patients, the sample size was still relatively small and therefore subject to bias. For example, the majority (54%) of patients who received single-agent anti-PD1/L1 were 65 years or older, whereas the majority of patients who received anti-PD1 + anti-CTLA4 (67%) were < 65 years old. Such unbalance in the patient population may reflect a treatment selection bias by the treating physicians. Another limitation was a lack of specialist central review of imaging studies. Response was defined based on investigator interpretation of radiological evidence showing tumor shrinkage and no new metastatic sites. As such, the definition of response did not adhere to standard RECIST criteria of 30% shrinkage in target lesion and could account for higher than expected response rates. The difference in the incidence of *BAP1* mutations between the anti-PD1/L1 and the anti-PD1 + anti-CTLA4 group is relevant. *BAP1* is a multifunctional tumor suppressor and *BAP1* mutations in UM are associated with activation of regulatory immune cells and an immunosuppressive tumor microenvironment [45, 46]. This could contribute to resistance to ICI and differences in clinical outcomes between our cohort; however, this is not well understood and is still being elucidated. Genomic analysis was not performed in the majority of patients included in this analysis; therefore, we did not assess whether the inclusion of genomic variable would provide additional prognostic value to our model. We were unable to evaluate the relationship between genomic differences between responders and non-responders as not all patients received exome sequencing.

## Conclusions

In summary, we present a novel prognostic model (MUMPS), comprised of three readily available clinical parameters, which can stratify mUM patients according to their expected prognosis and may have value in predicting benefit from specific ICI regimens. This study provides essential information for health care providers regarding the risk-benefit profile of anti-PD1 + anti-CTLA4. External validation of these data is warranted prior to clinical implementation.

## Data Availability

Availability of data and material: De-identified patient data are available upon reasonable request.

## DECLARATIONS

### Ethics approval and consent to participate

This study was approved by the University Health Network Institutional Research Ethics Board (REB# 19-5186).

### Patient consent for publication

Not required.

### Availability of data and material

De-identified patient data are available upon reasonable request.

### Competing interests

A.A.N.R.: An immediate family member is employed by Merck.

### Consultant/Advisory Board

A.A.N.R.: Pfizer; A.S.: Merck, Bristol-Myers Squibb, Novartis, Oncorus, Janssen. **Grant/Research support from Clinical Trials:** A.S.: Novartis, Bristol-Myers Squibb, Symphogen AstraZeneca/Medimmune, Merck, Bayer, Surface Oncology, Northern Biologics, Janssen Oncology/Johnson & Johnson, Roche, Regeneron, Alkermes, Array Biopharma. S.D.S.

## Funding

This research has been supported by research funds of A.S.

## Authors’ contributions

*D. Kelly and A.A.N. Rose contributed equally to this manuscript.

### Conception and design of study

D. Kelly, A.A.N. Rose and A. Spreafico

### Acquisition of Data

D. Kelly, A.A.N Rose, T.P. Muniz, D.W. Hogg, M.O. Butler, S. Saibil, J. Waldron, N. Laperriere, H. Krema, A. Spreafico

### Analysis and interpretation of data

D. Kelly, A.A.N Rose and A. Spreafico

### Writing and review of manuscript

D. Kelly, A.A.N Rose, T.P. Muniz, D.W. Hogg, M.O. Butler, S. Saibil, M. Iafolla, D.V. Araujo, J. Waldron, N. Laperriere, H. Krema, A. Spreafico ***Acknowledgements:*** A.A.N.R. is a recipient of a Hold ‘em For Life Clinician Scientist Award and a Douglas Wright Melanoma Award. T.P.M. and A.A.N.R. acknowledge fellowship funding from Alamos Gold Inc.

## Supplemental Figures

**Figure S1.**
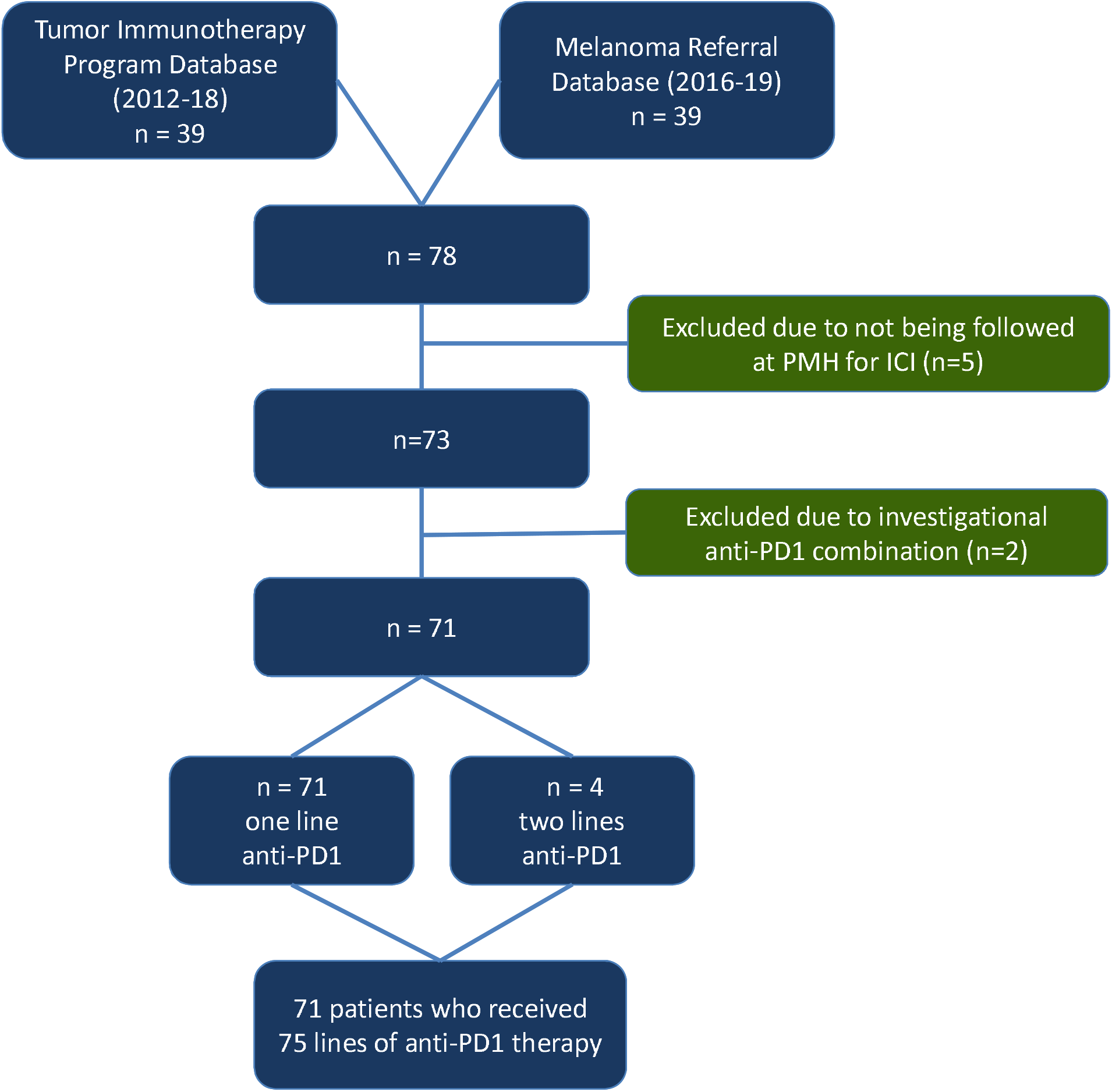

## Notes

### Author Declarations

DECLARATIONS Ethics approval and consent to participate: This study was approved by the University Health Network Institutional Research Ethics Board (REB# 19-5186). Patient consent for publication: Not required.

