## Supplemental Tables for "Development of a Metastatic Uveal Melanoma Prognostic risk Score (MUMPS) for use in patients receiving immune checkpoint inhibitors"

**Table S1: Treatment duration and discontinuation in the entire study cohort by treatment groups.**

|  | Anti-PD1  (n=54) | Anti-PD1+anti-CTLA4  (n=21) | P-value |
| --- | --- | --- | --- |
| Total # cycles Anti-PD1+anti-CTLA4 |  |  | - |
| 1 | - | 1 (5%) |  |
| 2 | - | 6 (29%) |  |
| 3 | - | 7 (33%) |  |
| 4 | - | 7 (33%) |  |
| Maintenance/Monotherapy |  |  |  |
| Anti-PD1 | 52 (96%) | 9 (43%) | P<0.0001 |
| Anti-PDL1 | 2 (4%) | 0 (0%) |  |
| No maintenance/monotherapy | - | 12 (57%) |  |
| # cycles containing standard dose anti-PD1 |  |  | P=0.0079 |
| Average | 7.2 | 3.5 |  |
| Median | 4 | 2 |  |
| Range | 1-38 | 0-29 |  |
| Reason for discontinuation |  |  | P=0.211 |
| Progression | 46 (85%) | 15 (71%) |  |
| Toxicity | 5 (9%) | 4 (19%) |  |
| On-going treatment | 1 (2%) | 2 (9%) |  |
| Lost to follow-up | 2 (4%) | 0 (0%) |  |

**Table S2: Treatment and dosing schedule of combination anti-PD-1 + anti-CTLA4**

| **Combinations** | **Number of patients**  *n*=21 (%) |
| --- | --- |
| anti-CTLA4 3 mg/kg + anti-PD1 1 mg/kg Q3W, followed by anti-PD1 3 mg/kg Q2W | 17 (81%) |
| Anti-CTLA4 1 mg/kg + anti-PD1 2mg/kg or 200 mg Q3W, followed by anti-PD1 2 mg/kg Q3W | 2 (9.5%) |
| anti-CTLA4 1 mg/kg + anti-PD1 3 mg/kg Q3W, followed by anti-PD1 3 mg/kg Q2W | 2 (9.5%) |

Abbreviations: Q2W = every 2 weeks; Q3W = every 3 weeks.

**Table S3: Genomic analyses of mUM tumor specimens**

|  | Entire cohort  (n=75) | Anti-PD1  (n=54) | Anti-PD1 +anti-CTLA4  (n=21) | P-value |
| --- | --- | --- | --- | --- |
| Impact Genetics |  |  |  | 0.128 |
| Yes | 16 (21%) | 9 (17%) | 7 (33%) |  |
| No | 59 (79%) | 45 (63%) | 14 (67%) |  |
| NGS |  |  |  | **0.032** |
| Yes | 50 (67%) | 32 (59%) | 18 (86%) |  |
| No | 25 (33%) | 22 (41%) | 3 (14%) |  |
| Impact Genetics |  |  |  |  |
| Chromosome 1 |  |  |  | 0.163 |
| 1p disomy | 9 (12%) | 5 (9%) | 4 (19%) |  |
| 1p borderline loss | 3 (4%) | 1 (2%) | 2 (10%) |  |
| 1p loss | 3 (4%) | 2 (4%) | 1 (5%) |  |
| Not tested/insufficient | 60 (80%) | 46 (85%) | 14 (67%) |  |
| Chromosome 3 |  |  |  | 0.170 |
| Monosomy 3 | 15 (20%) | 8 (15%) | 7 (33%) |  |
| Partial Monosomy | 1 (1%) | 1 (2%) | 0 (0%) |  |
| Not tested/insufficient | 59 (79%) | 45 (83%) | 14 (67%) |  |
| Chromosome 6p |  |  |  | **0.049** |
| 6p disomy | 12 (16%) | 5 (9%) | 7 (33%) |  |
| 6p gain/borderline gain | 3 (4%) | 3 (6%) | 0 (0%) |  |
| Not tested/ insufficient | 60 (80%) | 46 (85%) | 14 (67%) |  |
| Chromosome 6q |  |  |  | 0.212 |
| 6q disomy | 7 (9%) | 4 (74%) | 3 (14%) |  |
| 6q borderline loss | 2 (3%) | 1 (2%) | 1 (5%) |  |
| 6q loss | 6 (8%) | 3 (6%) | 3 (14%) |  |
| Not tested/insufficient | 60 (80%) | 46 (85%) | 14 (67%) |  |
| Chromosome 8p |  |  |  | 0.130 |
| 8p disomy | 8 (11%) | 6 (11%) | 2 (10%) |  |
| 8p borderline loss | 3 (4%) | 1 (2%) | 2 (10%) |  |
| 8p loss | 5 (7%) | 2 (4%) | 3 (14%) |  |
| Not tested/insufficient | 59 (79%) | 45 (83%) | 14 (67%) |  |
| Chromosome 8q |  |  |  | 0.164 |
| 8q gain | 14 (19%) | 7 (13%) | 7 (33%) |  |
| 8q partial gain | 2 (3%) | 2 (4%) | 0 |  |
| Not tested/insufficient | 59 (79%) | 45 (83%) | 14 (67%) |  |
| NGS Testing: |  |  |  |  |
| BRAF |  |  |  | 0.106 |
| BRAF WT | 49 (65%) | 32 (59%) | 17 (81%) |  |
| BRAF mutated | 0 | 0 | 0 |  |
| Not tested/insufficient | 26 (33%) | 22 (41%) | 4 (19%) |  |
| NRAS |  |  |  | 0.119 |
| NRAS WT | 46 (61%) | 30 (56%) | 16 (76%) |  |
| NRAS mutated | 0 | 0 | 0 |  |
| Not tested/insufficient | 29 (39%) | 24 (44%) | 5 (24%) |  |
| GNA11 |  |  |  | 0.088 |
| GNA11 209 | 15 (20%) | 11 (20%) | 4 (19%) |  |
| GNA11 183 | 2 (3%) | 2 (4%) | 0 |  |
| GNA11 WT | 27 (36%) | 15 (28%) | 12 (57%) |  |
| Not tested/insufficient | 31 (41%) | 26 (48%) | 5 (24%) |  |
| GNAQ |  |  |  | **0.024** |
| GNAQ 209 | 21 (28%) | 10 (19%) | 11 (52%) |  |
| GNAQ 183 | 1 (1%) | 1 (2%) | 0 |  |
| GNAQ WT | 22 (29%) | 17 (31%) | 5 (24%) |  |
| Not tested/insufficient | 31 (41%) | 26 (48%) | 5 (24%) |  |
| BAP1 |  |  |  | **0.003** |
| BAP1 mutated | 19 (25%) | 8 (15%) | 11 (54%) |  |
| BAP1 WT | 20 (27%) | 15 (28%) | 5 (24%) |  |
| Not tested/insufficient | 36 (48%) | 31 (57%) | 5 (24%) |  |
| SF3B1 |  |  |  | 0.062 |
| SF3B1 mutated | 6 (8%) | 4 (7%) | 2 (10%) |  |
| SF3B1 WT | 32 (43%) | 19 (35%) | 13 (62%) |  |
| Not tested/insufficient | 37 (49%) | 31 (57%) | 6 (28%) |  |

**Supplementary Table S4: Number of patients who received anti-PD1/L1 monotherapy or anti-PD1/L1 + anti-CTLA4 antibodies.**

| Treatment | MUMPS good risk  (score = 3) | MUMPS intermediate risk  (score = 2) | MUMPS poor risk  (score = 0 or 1) | P-value |
| --- | --- | --- | --- | --- |
| Anti-PD1 monotherapy | 17 | 28 | 9 | P=0.522 |
| Anti-PD1 + anti-CTLA4 | 5 | 10 | 6 |  |

**Supplementary Table S5: Open Interventional Clinical Trials for the treatment of Metastatic Uveal Melanoma (Current as of 24 January 2021)[1]**

| **Clinical Trial ID** | **Intervention** | **Phase** | **Size^a^** | **First Posted** |
| --- | --- | --- | --- | --- |
| NCT04589832 | Procaspase-Activating Compound 1 (PAC-1) in combination with Entrectinib (NTRK Inhibitor) | 1b/2 | 38 | Oct 2020 |
| NCT04552223 | Nivolumab and Relatlimab (LAG-3 Abtibody) | 2 | 27 | Sept 2020 |
| NCT04551352 | O7293583, A Tyrosinase-Related Protein 1(TYRP 1) -Targeting CD3 T-Cell Engager | 1 | 310 | Sept 2020 |
| NCT04335890 | Vaccination with Inhibitor Of Nuclear Factor Kappa B Kinase Subunit Beta (IKKb) matured Dendritic Cells | 1 | 12 | April 2020 |
| NCT04283890 | Percutaneous hepatic perfusion with Ipilimumab and Nivolumab | 1/2 | 88 | Feb 2020 |
| NCT04109456 | IN10018, a Focal Adhesion Kinase (FAK) Inhibitor +/-Cobimetinib | 1b | 52 | Sept 2019 |
| NCT03947385 | IDE196, a PKC inhibitor+/- Binimetinib in patients with solid tumors harboring GNAQ or GNAQ/11 mutations or PRKC fusions | 1/2 | 217 | May 2019 |
| NCT03865212 | Intravenous Injection of Vesicular Stomatitis Virus Expressing Human Interferon Beta and Tyrosinase Related Protein 1 (VSV-IFNb-TYRP1) | 1 | 71 | Mar 2019 |
| NCT03467516 | Tumor Infiltrating Lymphocytes | 2 | 59 | Mar 2018 |
| NCT03472586 | Immunoembolization in combination with ipilimumab and nivolumab | 2 | 35 | Mar 2018 |
| NCT03025256 | Intrathecal nivolumab in combination with intravenous Nivolumab in treating patients with leptomeningeal disease | 1 | 50 | Jan 2017 |
| NCT03068624 | Cyclophosphamide, autologous CD8+ SLC45A2-specific T lymphocytes via hepatic arterial infusion and ipilimumab | 1b | 30 | Mar 2017 |

^a^ Participant size as stated in registry entry

**Supplementary Figure Legends**

**Figure S1:** Consort Diagram. We identified 78 patients who had received anti-PD1/L1 containing therapy for metastatic uveal melanoma. Two patients were excluded due to non-uveal histology (blue nevus and conjunctival). Five patients were excluded who had received anti-PD1/L1 therapy combined with investigational agents (ie. not anti-CTLA4), Finally, we excluded who were not followed at PM-UHN during their ICI treatment. After exclusion of these patients, the remaining 67 patients received 1 line of anti-PD1/L1 and 4pts who had 2 lines of anti-PD1 ICI treatment. In total, we analysed data from 71 patients who had received a total of 75 lines of anti-PD1 ICI therapy.

1.	NIH, *Recruiting Studies ; Uveal Melanoma, Metastatic* NIH U.S. National Library of Medicine Clinical Trials.gov URL <https://clinicaltrials.gov/ct2/results?cond=Uveal+Melanoma%2C+Metastatic&Search=Apply&recrs=a&age_v=&gndr=&type=&rslt>= [Accessed 24 January 2021], 2021.
